## Supplemental table 1 for "REASSURED diagnostics at point-of-care in sub-Saharan Africa: A scoping review"

| Date of search | Database | Keywords | Search results |
| --- | --- | --- | --- |
| 26/07/2022 | SCOPUS | ( TITLE-ABS-KEY ( "point-of-care testing"  OR  "point-of-care testing"  OR  "Diagnostic Test"  OR  "point-of-care systems"  OR  "Point-of-Care Systems"  OR  "Diagnostic Test"  OR  point-of-care  OR  poct )  AND  TITLE-ABS-KEY ( "real time connectivity ease of specimen collection environmental friendliness affordable sensitive specific user friendly rapid equipment free delivered"  OR  reassured  OR  "REASSURED Devices"  OR  "REASSURED diagnostics"  OR  "REASSURED criteria" ) ) | 37 |
| 26/07/2022 | AFRICA-WIDE |  | 1 (grey literature) |
| 27/07/2022 | MEDLINE | ((MM "Diagnostic Test Kits Approval") OR (MM "Home Diagnostic Tests") OR (MM "Point-of-Care Testing") OR "point-of-care testing" OR "point-of-care testing" OR "Diagnostic Test" OR "point-of-care systems" OR "Point-of-Care Systems" OR "Diagnostic Test" OR Point-Of-Care OR POCT ) AND (real-time connectivity ease of specimen collection environmental friendliness affordable sensitive specific user-friendly rapid equipment-free delivered OR REASSURED OR REASSURED Devices OR REASSURED diagnostics OR REASSURED criteria) | 11 |
| 27/07/2022 | CINAHL | ((MM "Diagnostic Test Kits Approval") OR (MM "Home Diagnostic Tests") OR (MM "Point-of-Care Testing") OR "point-of-care testing" OR "point-of-care testing" OR "Diagnostic Test" OR "point-of-care systems" OR "Point-of-Care Systems" OR "Diagnostic Test" OR Point-Of-Care OR POCT ) AND (real-time connectivity ease of specimen collection environmental friendliness affordable sensitive specific user-friendly rapid equipment-free delivered OR REASSURED OR REASSURED Devices OR REASSURED diagnostics OR REASSURED criteria) | 6 |
| 14/07/2022 | DIMENSIONS | “Reassured diagnostics” | 141 |
| 28/08/2022 | ProQuest Central | ( TITLE-ABS-KEY ( "point-of-care testing"  OR  "point-of-care testing"  OR  "Diagnostic Test"  OR  "point-of-care systems"  OR  "Point-of-Care Systems"  OR  "Diagnostic Test"  OR  point-of-care  OR  poct )  AND  TITLE-ABS-KEY ( "real time connectivity ease of specimen collection environmental friendliness affordable sensitive specific user friendly rapid equipment free delivered"  OR  reassured  OR  "REASSURED Devices"  OR  "REASSURED diagnostics"  OR  "REASSURED criteria" ) ) | 54 |
| 28/07/2022 | Google Scholar | "point-of-care testing"  OR "Diagnostic Test"  OR  "point-of-care systems"  OR   "real time connectivity ease of specimen collection environmental friendliness affordable sensitive specific user friendly rapid equipment free delivered"  OR  reassured  OR  "REASSURED Devices"  OR  "REASSURED diagnostics"  OR  "REASSURED criteria" | 159 |
