## Supplemental table 3 for "REASSURED diagnostics at point-of-care in sub-Saharan Africa: A scoping review"

**S3 Table. Reviewers level of agreement: full text screening (Yes = 1, No = 0)**

| Author and Date | Reviewer 1 (BM) | Reviewer 2 (BL) |
| --- | --- | --- |
| Alarcon-Angeles 2022 | 0 | 0 |
| Bernabe-Ortiz 2021 | 1 | 1 |
| Calabria 2021 | 0 | 0 |
| Chua 2022 | 0 | 0 |
| Dexter 2022 | 1 | 1 |
| Diego 2022 | 0 | 0 |
| Gildner 2021 | 0 | 0 |
| Han 2020 | 0 | 0 |
| Hosseinifard 2021 | 0 | 0 |
| Hu 2022 | 0 | 0 |
| Ilyas 2020 | 1 | 0 |
| Juan 2022 | 0 | 0 |
| Kopkin 2022 | 0 | 0 |
| Land 2019 | 1 | 1 |
| Macchia 2022 | 0 | 0 |
| Murtagh 2021 | 0 | 1 |
| Park 2022 | 0 | 0 |
| Piorino 2022 | 0 | 1 |
| Schaumburg 2022 | 0 | 0 |
| Smith 2021 | 1 | 0 |
| Smith 2020 | 1 | 1 |
| Torne-Morato 2022 | 0 | 0 |
| Turbe 2021 | 1 | 1 |
| Kimani 2017 | 1 | 0 |
| Valera | 1 | 0 |

kap Reviewer1BM Reviewer2BL

| Expected agreement | Agreement | Kappa | Std. Err. | Z | Prob > Z |
| --- | --- | --- | --- | --- | --- |
| 76.00% | 56.16% | 0.4526 | 0.1966 | 2.30 | 0.0107 |

mcc Reviewer1BM Reviewer2BL

| Controls |

Cases	|	Exposed	Unexposed |	Total

-----------------+------------------------+------------

Exposed	|	5	4 |	9

Unexposed	|	2	14 |	16

-----------------+------------------------+------------

Total	|	7	18 |	25

McNemar's chi2(1) =	0.67	Prob > chi2 = 0.4142

Exact McNemar significance probability	= 0.6875

Proportion with factor

Cases	.36

Controls	.28	[95% Conf. Interval]

---------	--------------------

difference	.08	-.1494587	.3094587

ratio	1.285714	.7021954	2.354133

rel. diff. .1111111	-.1403524	.3625746

odds ratio	2	.2866338	22.1097	(exact)
